# Cognitive Processing Speed and Loneliness in Stroke Survivors: Insights from a Large-Scale Cohort Study

**DOI:** 10.1101/2023.10.09.23296730

**Authors:** Christopher Byrne, Rudi Coetzer, Richard Ramsey

**Affiliations:** School of Psychology & Sport Science, Bangor University, Bangor, Gwynedd, Wales, LL57 2AS, United Kingdom; Brainkind, 32 Market Place, Burgess Hill, West Sussex, RH15 9NP; Faculty of Medicine, Health & Life Sciences, Swansea University; Department of Health Sciences and Technology and Department of Humanities, Social and Political Sciences, ETH Zürich, Zürich, Switzerland

**Author notes:** Correspondence /.

**Keywords:** loneliness, stroke, acquired brain injury, British Cohort Study, cognition

## Abstract

**Objective:** Loneliness, when prolonged, is associated with many deleterious effects and has been shown to be highly prevalent in those with a history of stroke, yet the cognitive mechanisms underpinning this phenomenon remain unclear. Therefore, the current study aims to investigate the extent to which cognitive factors, with specific focus on processing speed, are associated with loneliness in those with a history of stroke.

**Method:** Utilizing data from the British Cohort Study (BCS), a nationally representative dataset, we conducted secondary data analysis. A total of 7,752 participants completed relevant questions related to health, social interactions, demographics, loneliness, and cognitive assessments. Among them, 47 had experienced a stroke (“stroke,” n = 47), 5,545 reported other health conditions (“ill,” n = 5,545), and 2,857 were deemed healthy (“healthy,” n = 2,857)

**Results:** Consistent with previous research, our findings confirmed a positive correlation between stroke history and heightened loneliness. However, inferential analysis revealed that processing speed, alongside other cognitive factors, had a minimal impact on loneliness, with correlations too small to draw definitive conclusions.

**Conclusion:** This study suggests that cognitive processing speed alone is not a robust predictor of loneliness in stroke survivors. Consequently, when developing interventions to combat loneliness in this population, it is crucial to consider a broader spectrum of factors, such as social engagement, emotional well-being, and interpersonal relationships. This underscores the imperative need for comprehensive assessments to better comprehend the multifaceted nature of loneliness and inform more effective intervention strategies.

## INTRODUCTION

Loneliness is defined as a discrepancy between the quality of one’s desired social relationships and one’s actual social relationships (Perlman & Peplau, 1998). Loneliness and social isolation, while theoretically connected, possess distinct conceptual, clinical, and empirical differences (Coyle & Dugan, 2012). Social isolation reflects objective aspects of social connection including frequency, duration, and proximity of others. In contrast, loneliness reflects a person’s subjective perception of the degree to which their social needs are being met. Therefore, being alone does not necessitate that one would experience loneliness. Equally, many people report experiencing loneliness despite being socially connected and integrated. As such, one can feel lonely in a crowd or find peace in solitude.

When loneliness is prolonged, it is associated with many deleterious effects both physiologically and psychologically, including poor sleep, anxiety, depression (Zawadzki, Graham & Gerin, 2013), high blood pressure and stroke (Hawkley, Thisted, Masi, & Cacioppo, 2010), as well as heart disease (Yanguas, Pinazo-Henandis, & Tarazona-Santabalbina, 2018). These substantial health effects are coupled with loneliness being experienced by 20-25% of the population at any one time (Cacioppo & Patrick, 2008; Smith & Kim, 2020), with more people becoming lonely in recent years (Groarke et al., 2020). Therefore, studying the causes and consequences of loneliness is a highly relevant topic of research, which spans the investigation of basic underlying mechanisms to real-world interventions.

The existing literature predominately investigates the causes and consequences of loneliness in older adult populations (Boss, Kang, & Branson, 2015) or those with neurodegenerative conditions, such as dementia (Victor, 2021). Little attention is given to those who have other non-progressive neurological conditions, such as Acquired Brain Injury (ABI). A specific sub-group of ABI that has been studied in the context of loneliness is stroke. For example, a recent large-scale study, which used nationally representative survey data in two separate samples of ∼8000 individuals, revealed that loneliness is highly prevalent in the stroke population, with up to one in three stroke survivors reporting higher levels of loneliness (Byrne, Saville, Coetzer & Ramsey, 2022). This initial work provided clear empirical evidence that loneliness was elevated in stroke survivors.

Theoretical papers, like Byrne, Salas, Coetzer & Ramsey (2022), have delineated how cognitive impairment post-brain injury could affect social connections, aligning with established loneliness models like the reaffiliation motive model (RAM; Qualter et al., 2015). For instance, damage to the left and right dorsolateral prefrontal cortex has been linked to deficits in executive control, involving challenges with planning, flexibility, and inhibition, crucial for socially adaptive behaviours (Moriguchi, 2014). Furthermore, Hertrich et al. (2021) discuss how damage to the left dorsolateral prefrontal cortex may lead to processing difficulties, potentially contributing to social isolation as individuals struggle with the complexity of social interactions. However, despite theoretical exploration in this area, there is limited empirical evidence demonstrating how cognitive mechanisms mediate the experience of loneliness following brain injury.

One of the most prevalent cognitive challenges faced by individuals with various forms of ABI is related to impairments in processing information efficiently and quickly, which constitutes the initial crucial step in encoding information into memory. Therefore, the primary aim of the current study was to estimate the extent to which loneliness varies as a function of processing speed. In addition, other cognitive factors, such as memory and verbal fluency, will be explored in both the healthy population and those with a history of stroke.

Stroke, ischaemic and haemorrhagic, is a leading cause of death and disability in the United Kingdom. The incidence rates for a first-time stroke are approximately 50 per 100,000 people. Thankfully, as medical interventions advance, there are more people surviving stroke each year. Approximately 1.3 million individuals are estimated to be living in the United Kingdom after experiencing a stroke (Stroke Association, 2021)). To provide some perspective, this figure signifies that roughly 2% of the UK population has a history of stroke. A significant portion of these individuals may experience enduring psychological, physical, and cognitive consequences, potentially rendering them susceptible to social isolation and loneliness (Stroke Association, 2021).

Recent work by Byrne and colleagues (2022) revealed that those with a history of stroke were at least 70% more likely to report loneliness when compared to a healthy population. In addition, the experience of loneliness in this population could not be explained by objective social factors such as frequency of social contact. This suggests that typical interventions addressing loneliness through increasing social contact alone may not be effective. Therefore, it is important to consider other non-social factors when considering alleviating loneliness in the stroke population. This may include examining whether the cognitive consequences of stroke impact on social functioning and, if so, what domains of cognition are important when considering the experience of loneliness in the stroke population.

To our knowledge, only a few studies have investigated the relationship between cognitive correlates and loneliness. The initial results in healthy populations are mixed, but generally show that loneliness is negatively correlated with several cognitive processes, such as processing speed, immediate memory, and delayed memory (Boss, Kang & Branson, 2015; Oakely & Deary, 2019). Impairments in executive functioning, including working memory and planning, have also been found to be a significant predictor of loneliness (Sin, Shao & Lee, 2021). Therefore, those that report more frequent experiences of loneliness tend to show lower performance in cognitive tasks.

Taking this a step further, Oakely & Deary (2019) showed that the results were not consistent across cognitive domains. For instance, processing speed, in addition to visuospatial ability, were more closely related to loneliness than other cognitive domains, such as verbal memory. This suggests that specific cognitive domains may be more crucial in mediating loneliness than others. These results are consistent with the proposal that some, but not all, cognitive impairments could disrupt an individual’s ability to engage in social interactions. Moreover, it suggests difficulties processing social cues (requiring processing speed ability), difficulty understanding and retaining the perspectives of others (requiring memory), and difficulty regulating their own emotions (requiring executive functioning), may all contribute to the experience of loneliness (Byrne, Salas, Ramsey & Coetzer, 2022).

The current study, therefore, aims to investigate the extent to which cognitive factors, with specific focus on processing speed, are associated with loneliness in those with a history of stroke. The study uses a large, secondary survey dataset from a nationally represented sample (British Cohort Study; BCS, N∼8581), which includes cognitive performance data, psychometric data related to loneliness and demographic information. Our pre-registered hypothesis was that loneliness scores will be associated with poorer cognitive functioning. More specifically, performance on cognitive tests measuring processing speed will be negatively correlated with loneliness scores. In addition, we hypothesised that this relationship will be stronger in those who report a history of stroke when compared to the healthy sample group.

## METHOD

### General methodological approach

The current study adopted a secondary data analysis approach using a large existing dataset collected via the British Cohort Study (BCS). The BCS is an ongoing multidisciplinary longitudinal birth cohort study that monitors over 17,000 individuals who were all born within a single week of each other in 1970 in England, Scotland and Wales. Participants involved in the BCS undertake surveys at specific time intervals collecting data relating to health and education, as well as social and economic variables. Surveys have been completed at the age of 5, 10, 16, 26, 30, 34, 38, 42 and more recently at ages 46-48. In the most recent survey (2019), variables of interest to the current study were included: cognitive performance indicators, measure of wellbeing (including indicators of loneliness), social contact, health status and demographic information. Further information and background regarding the BCS are provided by Elliott and Shepherd (2006). The primary analysis of interest was to estimate the relationship between a measure of cognitive performance (processing speed) and a measure of wellbeing that was closely associated with loneliness.

Since the BCS data does not include a validated and specific measure of loneliness, we used items from a measure of wellbeing as a proxy for loneliness. The wellbeing items on face value refer to themes that are closely associated with loneliness, such as feeling close to other people and feeling loved. Nonetheless, before running the primary analysis, we wanted to first empirically assess the extent to which these items correspond to a standardised measure of loneliness that has been specifically validated to index aspects of loneliness. As such, before the main analysis, we used a separate dataset (the National Survey of Wales [NSW]) to assess the extent to which the wellbeing items in question correspond to a measure of loneliness (The De Jong Gierveld Loneliness Scale; de Jong Gierveld & van Tilburg, 2006) in a large, nationally representative sample (∼8000 individuals). We report the findings from this preparatory analysis before completing the main analysis.

### Open Science Framework and Pre Registration

The BCS is freely available through the United Kingdom Data Services via UKdataservice.org. In addition, our analysis files, which outline each step of the current study’s analytical process, are provided on the open science framework (https://osf.io/rxgdy/) to facilitate transparency should other researchers wish to reproduce our analysis. Furthermore, the current study’s hypotheses, design and analyses were defined and preregistered in advance of data analysis (https://aspredicted.org/mu3iy.pdf). This promotes open science principles, recommended by Munafò, Nosek, Bishop et al (2017), and helps reduce questionable research practices such as HARKing (Hypothesising After Results are Known; Kerr, 1998).

### Deviation from pre-registration

The current study also aimed to examine the relationship between loneliness scores and biomarkers. It was hypothesised that there would be a strong positive relationship between inflammatory biomarkers and loneliness. However, it turned out that only 12 stroke participants completed both the wellbeing questionnaire items of interest and underwent the blood sample analysis that we would need to perform the biomarker analysis. Subsequently, we did not analyse the biomarker data any further as we felt that no meaningful inferences could be drawn from such a small sample. For transparency, we do plot the biomarker data in supplementary materials.

### Participants

A total of 8581 participants completed the most recent BCS survey (Age 46-48), with 7752 participants fully responding to questions related to variables of interest. Of the 8581 participants, 47 reported a history of stroke (“Stroke”; n = 47). The primary comparison groups of interest included those who reported any other form of health condition apart from stroke (“ill”; n = 5545) and those with no reported health difficulties (“Healthy”; n = 2857).

Unfortunately, 7 out of the 47 participants who reported a history of stroke did not answer the questions relating to *loneliness* leaving 40 post-stroke individuals in the final analysis. Similarly, only 2631 of the “Healthy” population and 5081 of the “ill” group answered questions relating to loneliness. The complete demographic characteristics, including sex, age, relationship status and socioeconomic status, for each group of interest is outlined in Table 1.

**Table 1.** Demographic information for Stroke, Healthy and Any Ill Health groups.

|  | Stroke | Healthy | Any Health Issue |
| --- | --- | --- | --- |
| **Gender** |  |  |  |
| Male | 25 (53.2%) | 1435 (50.2%) | 2632 (47.5%) |
| Female | 22 (46.8%) | 1422 (49.8%) | 2913 (52.5%) |
| **Age category** |  |  |  |
| 46 years old | 14 (29.8%) | 975 (34.1%) | 2168 (39.1%) |
| 47 years old | 24 (51.1%) | 1273 (44.6%) | 2297 (41.4%) |
| 48 years old | 9 (9.1%) | 609 (21.3%) | 1080 (19.5%) |
| **Marital Status\*** |  |  |  |
| Relationship/Married | 14 (31.1%) | 1792 (64%) | 3441 (63.3%) |
| No Partner | 30 (66.7%) | 998 (35.6%) | 1970 (36.2%) |
| Widowed Decile | 1 (2.2%) | 10 (0.4%) | 28 (0.5%) |
| **Level of Deprivation** |  |  |  |
| Most deprived 20% | 7 (14.9%) | 168 (5.9%) | 354 (6.4%) |
| 2<sup>†</sup> Decile | 6 (12.8%) | 165 (5.8%) | 404 (7.3%) |
| 3<sup>†</sup> Decile | 4 (8.5%) | 198 (6.9%) | 447 (8.1%) |
| 4<sup>†</sup> Decile | 4 (8.5%) | 244 (8.5%) | 472 (8.5%) |
| 5<sup>†</sup> Decile | 4 (8.5%) | 299 (10.5%) | 543 (9.8%) |
| 6<sup>†</sup> Decile | 7 (14.9%) | 304 (10.6%) | 606 (10.9%) |
| 7<sup>†</sup> Decile | 1 (2.1%) | 353 (12.4%) | 634 (11.4%) |
| 8<sup>†</sup> Decile | 4 (8.5%) | 361 (12.6%) | 681 (12.3%) |
| 9<sup>†</sup> Decile | 3 (6.4%) | 340 (11.9%) | 683 (12.3%) |
| Least deprived 20% | 7 (14.9%) | 420 (14.7%) | 704 (12.7%) |

\*45 stroke participants answered relationship status question
† Level of Deprivation in Deciles.

### Measures

### Dependent Variable/Outcome Variable

Loneliness was measured using three items within The Warwick-Edinburgh Mental Wellbeing Scales (WEMWBS). The WEMWBS is a 14-item questionnaire developed to enable the monitoring of mental wellbeing in the general population (Tennant, Hiller, Fishwick *et al.,* 2007). Although the WEMWBS is validated to measure overall wellbeing, and not exclusively loneliness, it includes three questions that relate closely to an individuals’ subjective experience of their social situation: 1) “*I’ve been feeling interested in other people*”; 2) “*I’ve been feeling close to other people*”, and; 3) “*I’ve been feeling loved”.* Each item is measured on a Likert scale from 1 – 5 (1-“none of the time”, 2 – “rarely”, 3 – “some of the time”, 4 – often, 5 – “all of the time”). For the purpose of the analysis, each item was reversed scored so that higher scores would reflect a proxy measure for greater levels of loneliness. As the three items of the WEMWBS have not been validated to measure loneliness specifically, further analyses of the items were performed to examine the suitability of use as a proxy measure for loneliness in the current study (see preparatory analysis section below for details).

### Independent Variables/Predictor Variables

#### Cognitive Factors

The most recent BCS also comprised of a small cognitive assessment battery measuring memory (immediate and delayed), verbal fluency (a common measure of executive functioning), and processing speed.

##### Processing Speed

Processing speed was measured using a letter cancelation task (LCT). The use of LCTs as a measure of processing speed is well documented in the neuropsychological literature (Haarhoff, Gadd, Semenya & Van Eeden, 2020). In the BCS, participants were presented with a page of randomised alphabetical letters in rows (n = 26) and columns (n = 30). The participants were then asked to cross the letters “P” and “W” as quickly as possible within a one-minute time limit. The BCS survey captured both the accuracy and speed of participants’ performance. The speed score is the variable of interest for the current study, which related to the total number of correct letters scanned within the time limit. Higher scores indicate faster processing speed. The decision to focus primarily on speed in our analysis was rooted in the specific research aims and hypotheses of our study. We recognise that both accuracy and speed are crucial dimensions when assessing cognitive performance clinically. However, for the purpose of our investigation, we chose to prioritize speed as the primary variable of interest. As the task only captured correctly identified letters, this ensured that the speed scores reflects efficient processing, rather than simply rapid scanning with many errors.

##### Verbal Fluency

Verbal fluency was assessed through an animal naming task. Animal fluency tasks are frequently used in clinical settings due to their ability assess semantic processing and verbal ability (Shao, Janse, Visser, & Meyer, 2014). Participants were asked to name as many different animals as possible within a one-minute time period. Repetition of the same animal, redundancies (e.g. white swan, black swan) and named animals (bugs bunny) were excluded from the score. Higher scores indicate better verbal fluency/executive control.

##### Immediate Memory & Delayed Memory

Immediate memory ability was assessed via a Verbal List Learning Task (10 words). Verbal List Learning Tasks are often routinely employed in neuropsychological assessment as tool to examine encoding, storage, and retrieval (Rabin, Paolillo & Barr, 2016). Participants were presented with a list of 10 words through a computer using a pre-recorded voice. If the computer was deemed inaudible to the participant, then the interviewer would read the list aloud imitating the pace of the pre-recorded voice at a rate of one word every two-second. Once the list had been presented, the participants had 2 minutes to recall as many words as they could remember. For the delayed memory phase, the participants were required to recall words from the initial 10-word list following a distraction period consisting of verbal fluency and processing speed tests. Again, participants were given a time limit of 2 minutes to recall as many words as they could remember in the delayed memory phase. The time between the initial presentation of the 10 words and the delayed recall task was approximately 5 minutes.

#### Health, demographic and social contact variables (covariates)

##### Health status

The BCS included items investigating health status. Participants were initially asked an open-ended question relating to if they have any longstanding physical or mental health conditions - “*Do you have any physical or mental health conditions or illnesses lasting or expected to last 12 months or more?”.* If yes, the participants were asked specifically to choose which health conditions apply from a pre-defined selection – “*have you had any of the health problems listed on this card? Please include any health problems that had already started before that date. You can tell me which numbers apply*”. Participants who selected “Stroke” out of the pre-defined medical conditions were allocated to the stroke group. For the purpose of the current study, two comparison groups were also selected (“Healthy” & “Ill Health”) for comparison to the group of interest (“Stroke”). The “Ill Health” group included all participant who reported “yes” to the initial health screening item, with exception of those who also selected stroke. The “Healthy” group included all participants who reported “no” to the initial health screening question.

##### Demographic factors

Demographic factors captured by the BCS were also included in the analysis such as gender (binary; Male or Female), age (continuous; 46-48 years), relationship status & socio-economic deprivation (Index of Multiple Deprivation (IMD)). The IMD is a measure of relative deprivation in small areas in the UK. It is calculated by combining data from a number of different sources, including income, employment, education, health, crime, barriers to housing and services, and living environment. The IMD ranks geographical areas into deciles: 1^st^ decile (most deprived area) to 10^th^ decile (least deprived area). Relationship status was captured by the BCS included: 1. Separated, 2. Married, 3. Divorced, 4. Widowed, 5. Civil partner, 6. Former civil partner, 7. Surviving civil partner, & 8. Never married or in civil partnership/Single. The current study further categorised relationship status into three groups: Single, Married or Widowed.

##### Frequency of social contact

Two items of the BCS examined frequency of social contact with friends and family: Item 1 – contact with family (“how often do you meet with family”), Item 2 – contact with friends (“how often do you meet with friends”). Likert scale response range from 1 (“three of more times a week”) to 7 (“never”). These measures are used by the current study to examine objective social isolation.

### Analyses

#### Preparatory analyses of NSW WEMWBS data

We wanted to assess the extent to which there is an association between the three WEMWBS loneliness related items and another well-established measures of loneliness, the De Jong Gierveld Loneliness Scale (de Jong Gierveld & van Tilburg, 2006). To do this, data from the National Survey of Wales (NSW 2016/17) were used. The NSW2016/17 is a survey involving 10,493 participants from a nationally represented sample of an entire country of Wales. Details of this survey can be found in the Welsh Government Technical Report by Helme & Brown (2018). Pertinent to the current study, the NSW2016/17 included both the WEMWBS and De Jong Gierveld Loneliness Scale. This allowed for additional analyses to test for the degree of correspondence between the two measures.

We performed three main analyses to estimate the correspondence between the DJG loneliness scale and the three items of the WEMWBS. Overall, the aim of this preparatory analytical stage was to provide some confidence that there is reasonable correspondence between the three WEMWBS loneliness items and the De Jong Gierveld Loneliness Scale from a data-driven perspective. From a conceptual perspective, it is clear that there is similarity between the different scales and that they all relate to key features of loneliness. However, the empirical demonstration of data-driven similarity within a large, nationally representative sample would provide further support that the WEMWBS loneliness items can be used as a proxy for a validated measure of loneliness. In other words, if our primary dataset did include the De Jong Gierveld Loneliness Scale, we would have expected to observe similar results.

Although we find this preparatory step valuable and an important justification for using the WEMWBS items in the main analysis, we do acknowledge that this analytical approach does not represent a formal validation of the WEMWBS items as measures of loneliness specifically. A formal validation paper would be a substantial piece of work in itself (Furr, 2022), and that is not the focus of our work here. As such, while we do think that the inferences that can be drawn from the primary analysis benefit in important ways from this preparatory analysis, we want to make clear that this is not a formal validation of the scales as measures of loneliness.

We briefly summarise each of the three preparatory analyses below. For full details of the preparatory analysis phase, including the analysis scripts, plots and tables, please see our OSF page (https://osf.io/rxgdy/) and supplementary materials.

First, we calculated the correlation strength between all four variables. If the DJG scale and WEMWBS items vary in a similar manner across the sample, we would expect a positive correlation.

Second, for each item separately, we calculated the percentage of the sample that responded within each available category option. We then visualised and compared the distribution of these responses between the De Jong Gierveld Loneliness Scale and the three WEMWBS items. A similar distribution of responses between the scales would be suggestive that they are capturing something common in the way participants respond.

Third, we ran multiple regression analyses to estimate the extent to which variables predict WEMWBS items in the same way as De Jong Gierveld Loneliness Scale (DJGS). For example, we asked if there was a correspondence between how variables such as gender, social contact, health etc., predict a validated measure of loneliness, as well as the three WEMWBS items that relate to other people. If the factors predict WEMWBS items in a similar direction and strength as the De Jong Gierveld Loneliness Scale, then we would consider that as further evidence for a correspondence between the three WEMWBS items and the De Jong Gierveld Loneliness Scale.

#### Primary analysis of BCS data

We performed data analyses in the R programming language (v4.0.4; R Core Team, 2021). Our general analysis strategy followed a Bayesian estimation approach to multiple regression (McElreath, 2020). The basic logic of this approach is to estimate and evaluate parameters of interest across a series of increasingly complex models, as well as to perform model comparison between simpler and more complex models. More specifically, we followed a recent translation of McElreath’s (2020) general principles into a different set of tools (Kurz, 2020). As such, we use the Bayesian modelling package ‘brms’ to build regression models (Bürkner, 2017, 2018). Furthermore, we follow the ‘tidyverse’ principles (Wickham & Grolemund, 2016) and generate plots using the associated data plotting package ‘ggplot2’, as well as the ‘tidybayes’ package (Kay, 2020).

We first describe in detail our pre-registered analysis pipeline, which focussed on the relationship between processing speed, stroke and loneliness. For any further analyses, the same basic approach was also employed. Given that the dependent variable is an ordered category (a 1-5 rating scale), we used ordinal regression. We built multivariate models using the ‘mvbind’ function in brms, which means that we could include all three DVs in one model.

We calculated six multivariate models, which we built incrementally in complexity. Our model labelling convention was as follows: ‘b’ = bayesian; ‘p’ = processing speed; and then a number to denote the level of complexity. As such, model bp0 was an ‘intercepts only’ model, just so that we could compare subsequent models that included predictors of interest to a model without any predictors. Model bp1 additionally included demographic variables (gender, relationship status and deprivation), all of which were coded categorically. Model bp2 additionally included the frequency of social contact (friend and family frequency, both of which were centred and standardised). Model bp3 included processing speed, which was centred and standardised. Model bp4 included the health variable (heathy vs. stroke). Model bp5 included the interaction between health and processing speed and constituted the full model.

The brms formula for model bp5 is specified here:

brm(mvbind(people, close, loved) ∼ 1 + sex + relationship_status + deprivation + family_frequency + friend_frequency + cognitive_speed * health, family = cumulative(“probit”))

Note: ‘people’, ‘close’ and ‘loved’ refer to the three wemwbs scales that served as our dependent variables.

We set priors using a weakly informative approach (Gelman, 2006). Weakly informative priors differ from uniform priors by placing a constrained distribution on expected results rather leaving all results to be equally likely (i.e., uniform). They also differ from specific informative priors, which are far more precisely specified, because we currently do not have sufficient knowledge to place more specific constraints on what we expect to find. We used the probit link in our cumulative models, which means our priors are specified in a standardised metric (z-scores). We placed priors for the thresholds at zero with a normal distribution of 1.5. The fixed effects or predictors, as well as the standard deviations, were centred around zero with a normal distribution of 1. This means that we expect effects of interest (population effects) to be around zero more than we do 1 unit of standardised difference. Given that effects are relatively small in psychology, we felt that this was reasonable expectation (Funder & Ozer, 2019).

We evaluated these models with regard to our hypotheses in two main ways. First, we evaluate the key parameters of interest from the posterior distribution of the full model. Therefore, we primarily evaluate the parameter estimate and distribution of interaction between processing speed and health in model bp5. Second, we performed model comparison via efficient approximate leave-one-out cross validation (LOO; Vehtari et al., 2017). LOO is a way of estimating how accurately the model can predict out-of-sample data. Therefore, we took all models and asked how accurate they were at predicting the out-of-sample data. By doing so, we can estimate how much increasing model complexity increases model accuracy.

Descriptive analyses were also performed to outline the characteristics of the data set for each variable of interest. Demographic characteristics for each group (‘stroke’, ‘healthy’ and ‘ill health’) including age, gender, marital status and socioeconomic deprivation is outlined in *Table 1*. To examine whether there were any mean differences between cognitive performance scores across groups a multivariate analysis of variance was completed. The results of this analysis are outlined in *Table 2.* A descriptive analysis was then completed to examine loneliness scores across all groups for each loneliness domain (‘closeness’, ‘interest’ and ‘loved’) which is outlined in *Table 3*. In order to examine whether those with a history of stroke report higher levels of loneliness compared the ‘ill health’ and ‘healthy’ comparison groups, mean differences and confidence intervals (95%) were generated for each loneliness item. Based on previous research (Byrne et al, 2022), we expected the stroke group to report higher levels of loneliness compared to both comparison groups.

**Table 2.** Mean difference in performance across four cognitive tests for the Stroke, Healthy and Any Ill Health groups.

| Cognitive test | Health Status | Health Status | Mean difference | Std. Error | *p* value | 95% lower CI | 95% Upper CI |
| --- | --- | --- | --- | --- | --- | --- | --- |
| Processing speed | Stroke | Ill Health | -38.132* | 13.145 | .011 | -69.61 | -6.66 |
|  |  | Healthy | -38.294* | 13.195 | .011 | -69.89 | -6.70 |
| Verbal Fluency | Stroke | Ill health | -4.142* | .917 | .000 | -6.34 | -1.95 |
|  |  | Healthy | -3.931* | .920 | .000 | -6.14 | -1.73 |
| Immediate Memory | Stroke | Ill Health | -.925* | .212 | .000 | -1.43 | -.42 |
|  |  | Healthy | -.914* | .213 | .000 | -1.42 | -.40 |
| Delayed Memory | Stroke | Ill Health | -1.046* | .271 | .000 | -1.70 | -.40 |
|  |  | Healthy | -1.068* | .272 | .000 | -1.72 | -.42 |

**Table 3.**
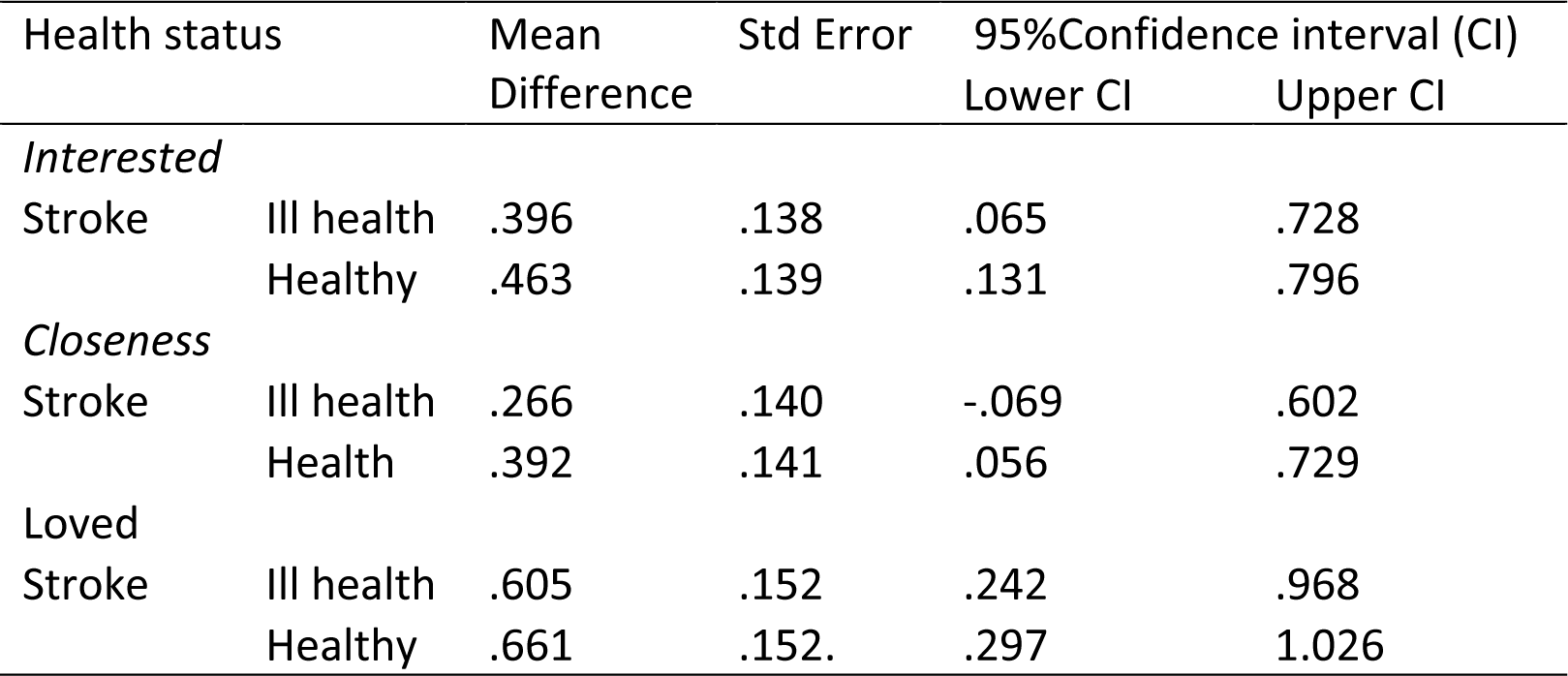
Mean difference between loneliness scores across groups.

The Bayesian approach taken offers a distinct framework for parameter estimation, one that inherently accounts for uncertainty and integrates prior information into the analysis process. Within this Bayesian paradigm, the need for conventional corrections for multiple comparisons is mitigated. This is outlined in the works of Gelman and colleagues (2000; 2012). Therefore, the decision not to correct for multiple comparisons stemmed from the fundamental distinctions in the estimation of parameters of interest between Bayesian and frequentist approaches.

## RESULTS

### Preparatory analyses of NSW WEMWBS data

We ran three different analyses to estimate the correspondence between the WEMWBS items of interest and the DJGS. First, a correlational analysis was completed demonstrating that the three items of the WEMWBS positively correlated with the DJGS with the correlation coefficients ranging from r = 0.3 to 0.65, but with most ∼ 0.5 or higher (Supplementary Figure 1). Second, we plotted the distribution of responses between the DJGS and the WEMWBS and they were found to be visually similar suggesting that there is a commonality between the two scales (Supplementary Figure 2). Third, a regression analysis was completed revealing that the same demographic variables used in the current study (gender, age etc.) predict both WEMWBS and DJGS scores in a similar manner, which again suggests that there is a commonality between the two scales (Supplementary Figure 3). Subsequently, although it is noted that the analyses completed do not reflect a more formal validation of the WEMWBS items as a loneliness measure, it does suggest good face validity and therefore increase our confidence in using it as a proxy measure of loneliness.

### Main analysis

#### Sample characteristics

Demographic characteristics for each sample group are outlined in Table 1. Demographic factors were mostly equal across groups except for relationship status. Those in the Stroke group (31.1%) were less likely to be in a relationship when compared to those in Healthy (64%) and Ill Health (63.3%) groups. Given the way the BCS was created, there was little difference in groups in age. Equally, there were little difference in social-economic deprivation between all three groups. However, a difference was observed for those in the most deprived areas, with 14.9% of the Stroke group fell within the most deprived decile, compared to 5.9% of the Healthy group and 6.4% of the ‘Any Ill Health’ group.

Difference between groups for cognitive performance were also examined. Those with a history of stroke performed more poorly across all cognitive domains including processing speed, immediate memory, delayed memory and verbal fluency, when compared to both the Health group and the Ill Health group.

#### Health conditions and loneliness

The mean difference and 95% confidence interval between the healthy, ill and stroke groups on each loneliness item are reported in Table 3 and plotted in Figure 1. For each item, the stroke group reported higher levels of loneliness than the healthy and otherwise ill groups. These findings are reassuring as they are similar to those reported previously, which showed elevated levels of loneliness in post-stroke individuals (Byrne, Saville et al., 2022).

**Figure 1.**
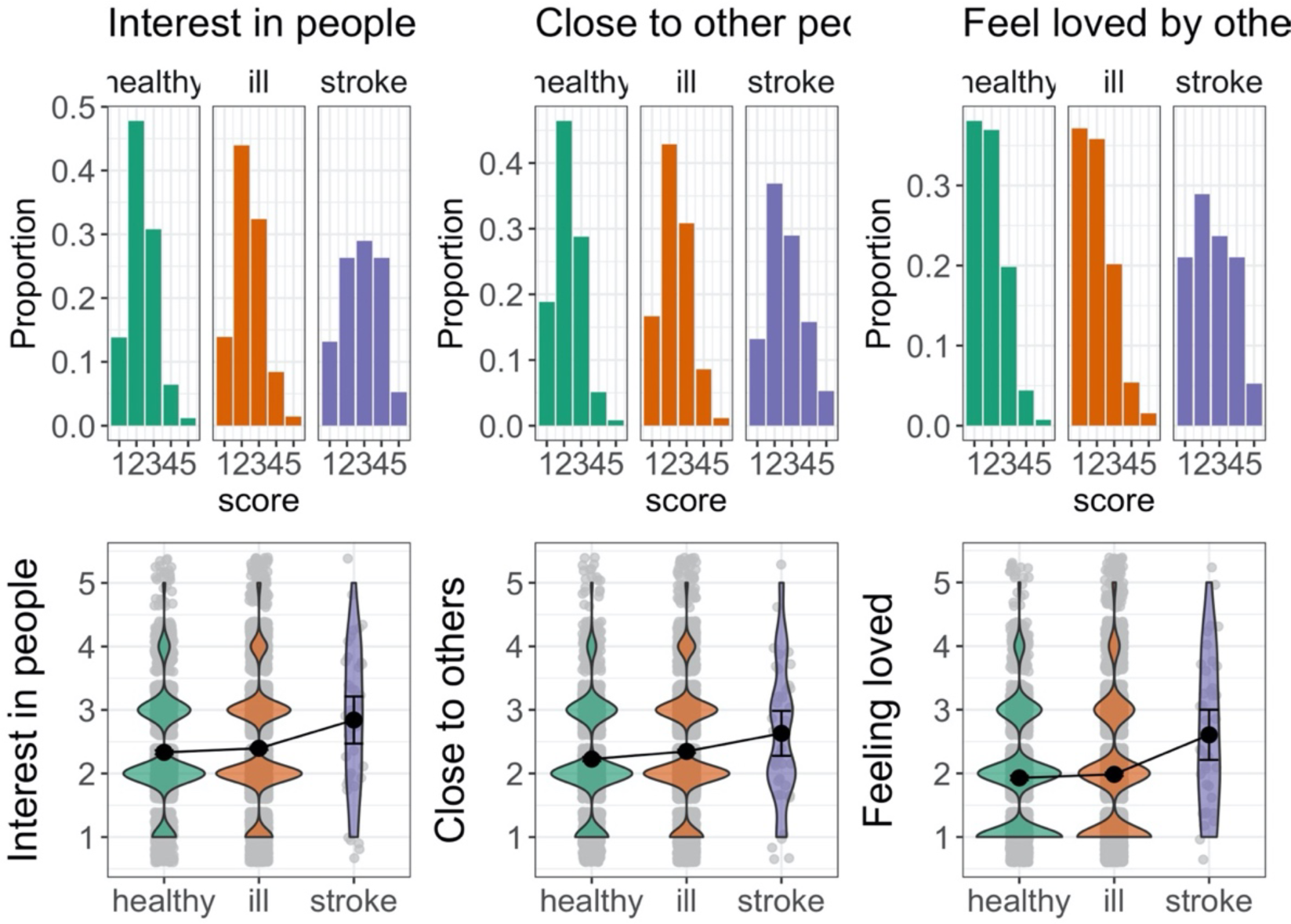
Histogram and violin plots demonstrating the distribution mean scores on the loneliness WEMWEBS items across all participant groups.

#### Cognitive correlates, loneliness and stroke

First, we assessed key parameters of interest in the full model (Figure 3; Table 4). The general pattern for our key parameters is the same for all three dependent variables (feeling loved, feeling interested, and feeling close).

**Figure 2.**
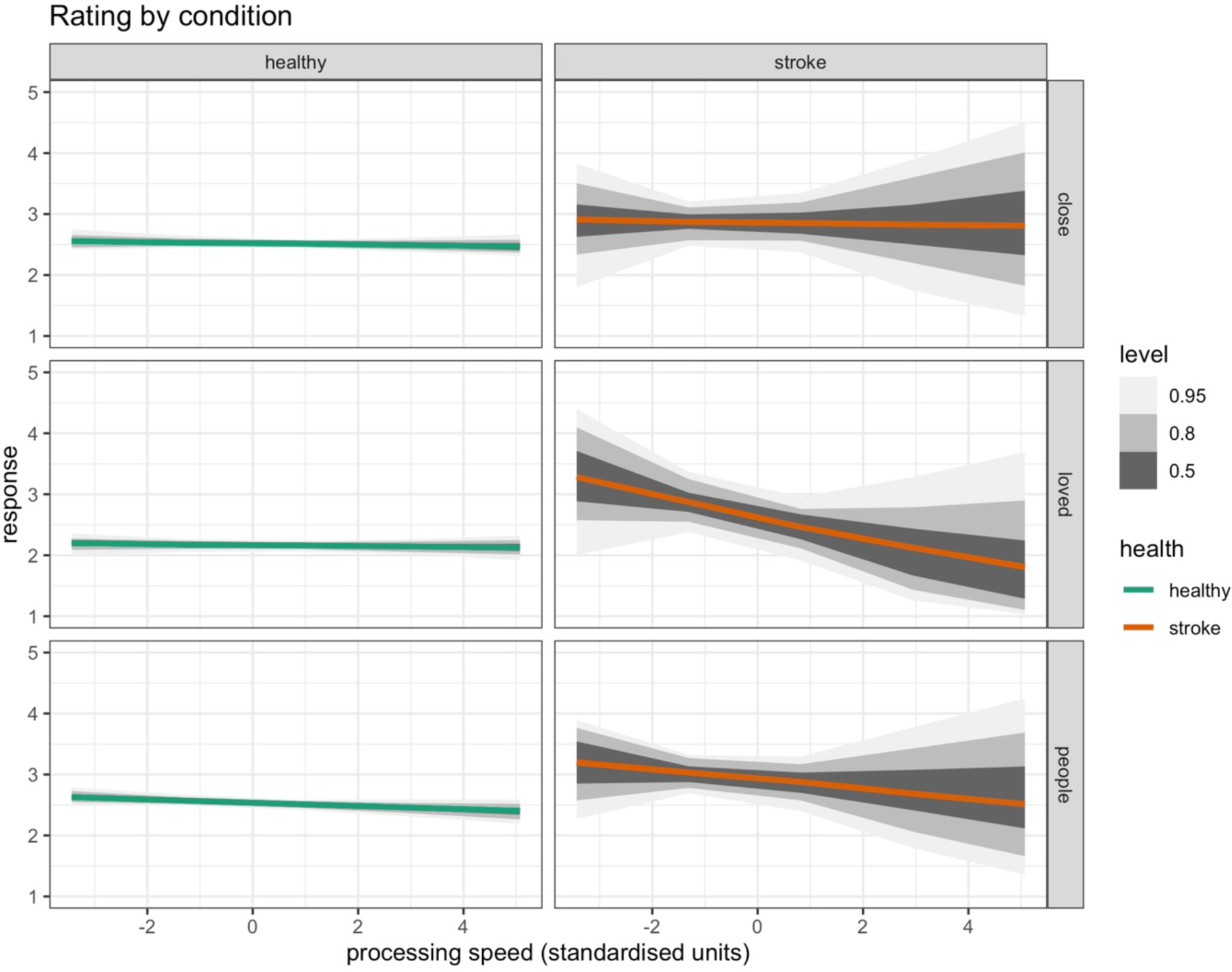
Coefficient slopes for loneliness and processing speed for healthy and stroke groups.

**Figure 3.**
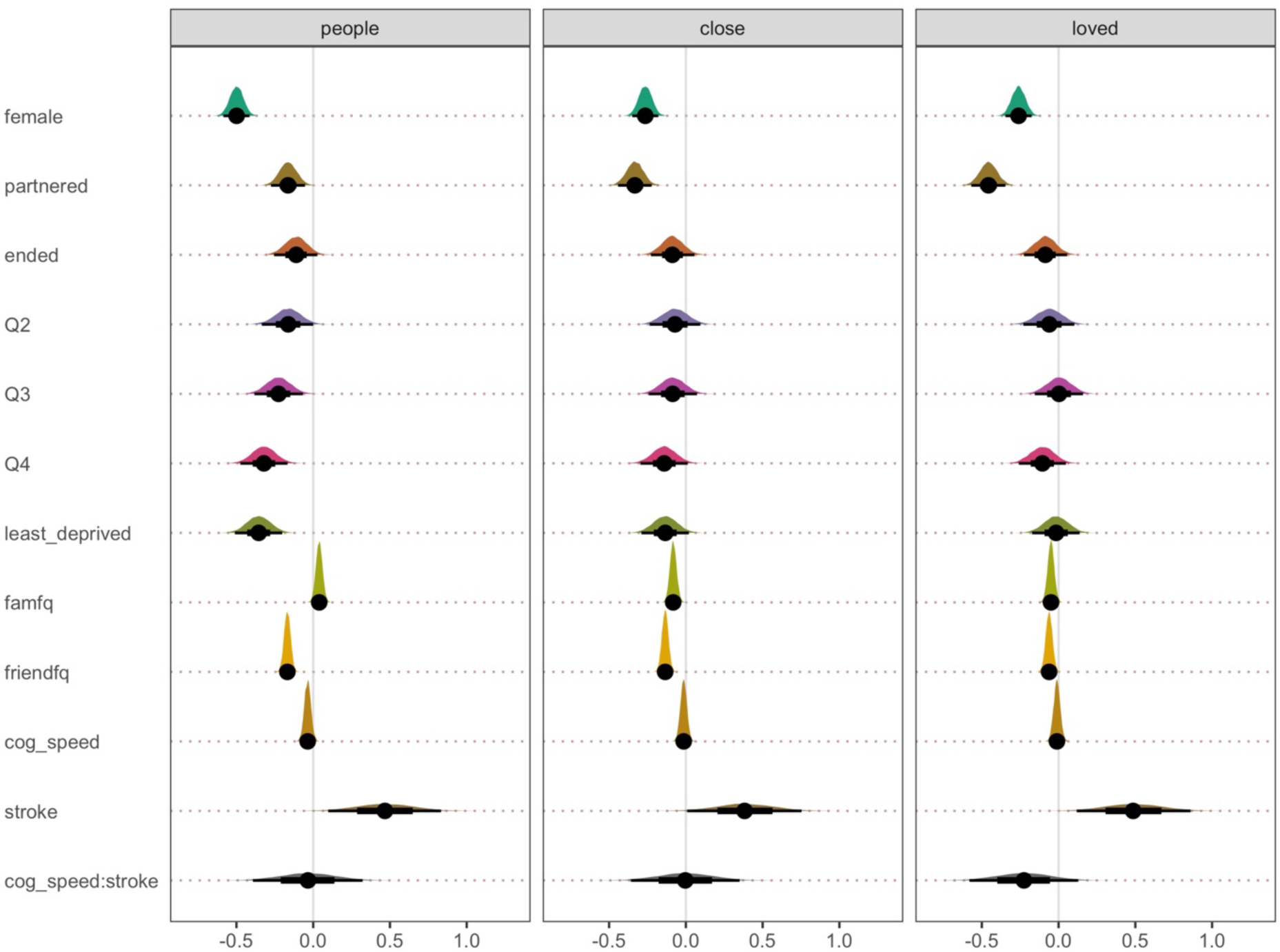
Multivariate model coefficient plot for fixed effects (predictors)

**Table 4.**
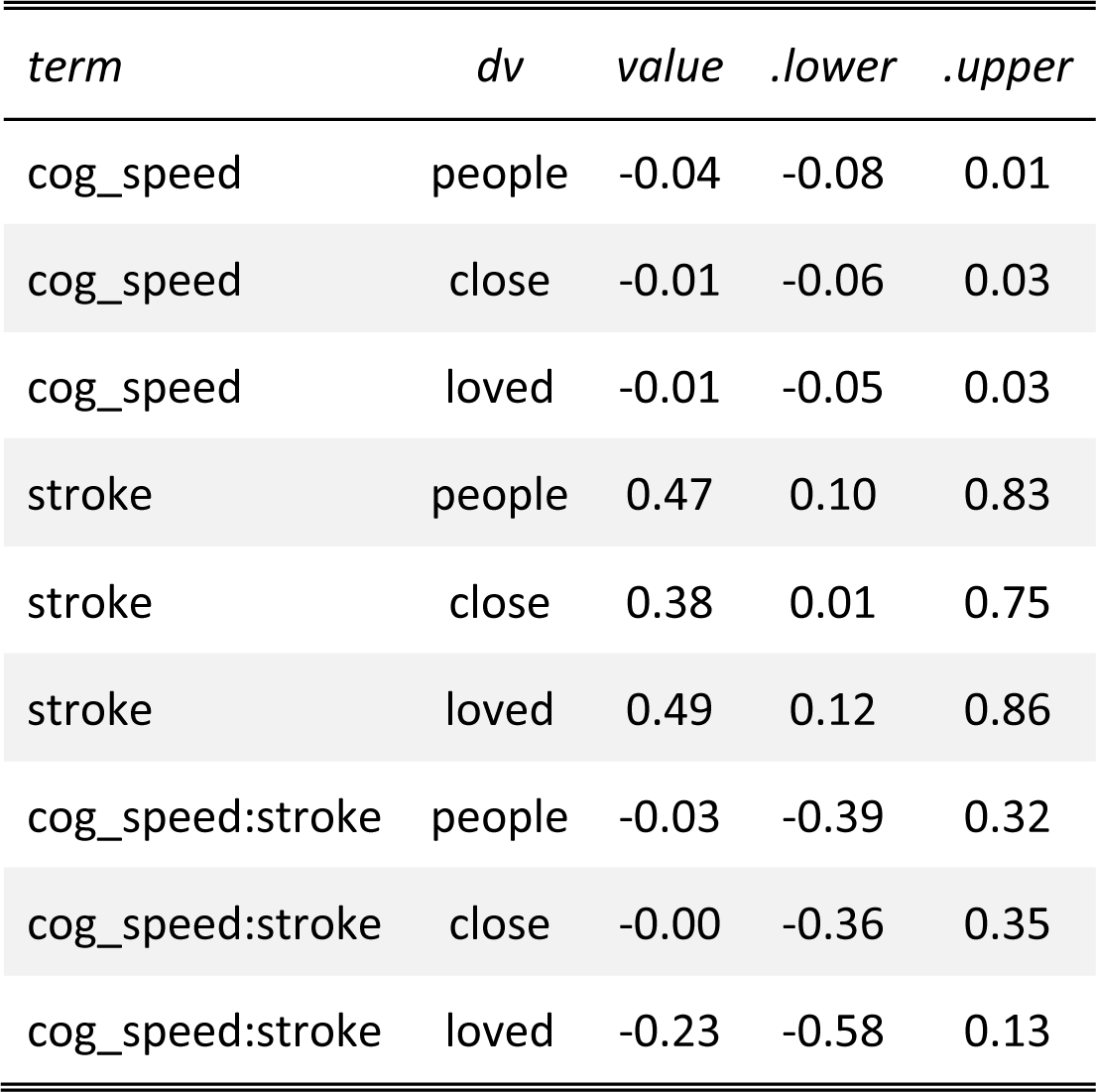
Parameter estimates for fixed effects.

We start by interpreting the processing speed and health group interaction, which was one of our key pre-registered effects. The posterior distribution for the interaction term shows considerable overlap with zero, which suggests that the relationship between processing speed and loneliness does not differ substantially between the healthy and stroke groups.

We then separately evaluated the effects of having a stroke and processing speed in relation to loneliness. Compared to the healthy group, having a stroke increased the degree of loneliness reported, which replicates prior work from our lab (Byrne et al., 2021). Stroke was associated with an increase in reported loneliness of 0.5 standardised units or an average of approximately 0.5 points on a scale from 1-5 (Figure 3).

In contrast, processing speed showed no clear and obvious relationship with loneliness (Figure 3). The posterior distribution for the average effect of processing speed was very close to zero and showed clear overlap with zero. This finding suggests that, in general, variations in cognitive processing speed do not play a substantial role in determining levels of loneliness across the entire sample.

Given that past research and our pre-registered hypotheses suggested that we should expect a negative relationship between processing speed and loneliness, such that those with faster cognitive processing speed should report lower levels of loneliness, we make a few additional observations. Visual inspection of the slopes in Figure 2 shows that in the healthy group, there was a very subtle negative slope, which is consistent with our expectations and past research. However, the relationship is too small to make any clear inferences about it. Furthermore, in the stroke group, the negative slope is steeper for “feeling loved” and the “interested in people” items. Such findings are numerically consistent with our pre-registered hypotheses that the relationship between processing speed and loneliness with be greater in the stroke group compared to the healthy group. However, as stated above, our modelling estimates remain too uncertain to form clear inferences about these relationships.

Second, we performed model comparison between simpler and more complex models. As can be seen in Figure 4, once predictors, such as demographic factors (bp1) and social contact (bp2), were added to the model, out-of-sample predictive accuracy improved. However, there was no clear difference between the models with predictors, which suggests that adding additional variables, such as cognitive processing speed (bp3) and health (bp4), did not make a substantial improvement, in terms of out-of-sample predictive accuracy.

**Figure 4.**
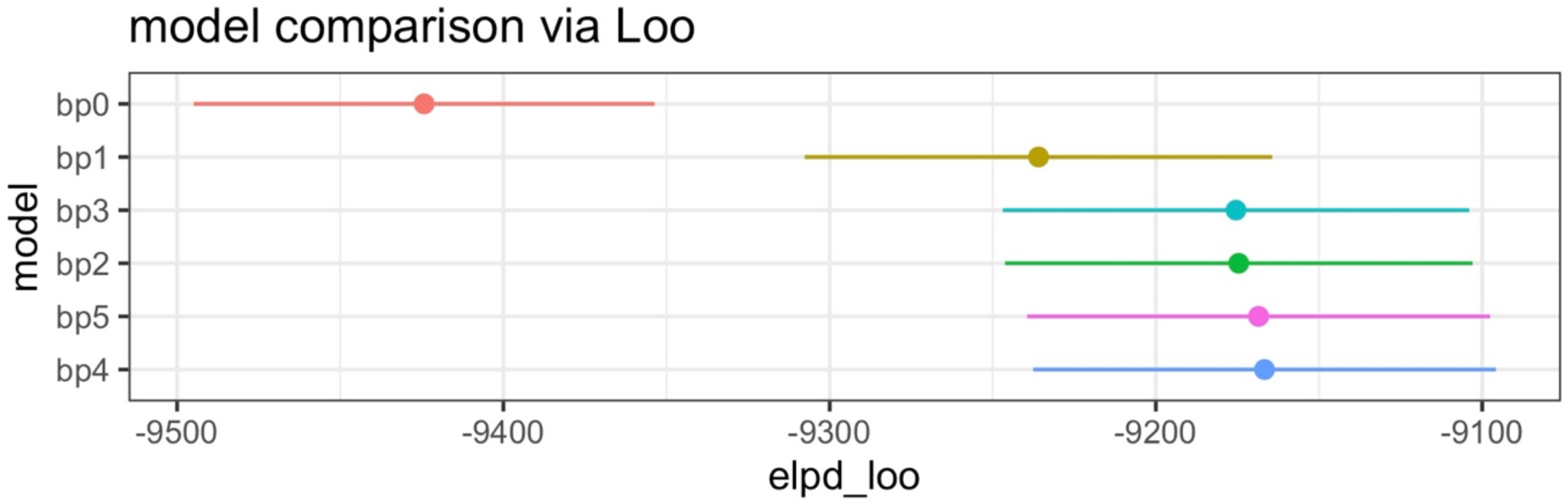
Visual representation of model comparison. Note. elpd_loo: estimate of the expected log pointwise predictive density; loo: leave-one-out estimated cross validation; error bars: standard error of the mean.

To further explore the relationship between other cognitive factors and loneliness, supplementary analyses were conducted to examine other cognitive domains such as immediate memory, delayed memory, and verbal fluency. Surprisingly, the additional analyses did not reveal any significant associations between these cognitive domains and loneliness in either the stroke or healthy group (see additional analyses on our OSF page).

## DISCUSSION

The present study sought to explore potential associations between processing speed, health status, and loneliness. In line with prior research (Byrne et al., 2022), we show that experiencing a stroke is positively correlated with heightened levels of loneliness. However, the primary analysis did not reveal any clear and obvious interactive effect between processing speed and health status (healthy or stroke), which indicates that the relationship between processing speed and loneliness is largely invariant to the presence of stroke. Consequently, our inferential analysis suggests that processing speed, as operationalised through a Letter Cancelation Task, has minimal influence on the experience of loneliness, and this association remains unaltered in the aftermath of a stroke.

Visual inspection of the data revealed a small negative slope in the healthy group, which is consistent with our pre-registered hypotheses and prior studies (Gilmour, 2011; Gow et al., 2013; O’Luanaigh et al. 2012; Sin, Shoa, & Lee, 2021; Boss et al. 2015). However, in contrast to other studies, such as Boss, Kang, & Branson, (2015) and Sin, Shao, & Lee, (2021) who demonstrated strong negative correlations, the size of the correlation in the current study is too small to draw clear inferences. Our findings are in line with Gow et al (2013) who also demonstrated a relationship between loneliness and processing speed, but the size of the effect was again small (Partial eta squared 0.009). The current study revealed a steeper negative slope in the stroke group for certain aspects of loneliness (feeling loved & feeling interested in people) when compared with the healthy population, which aligned with our hypotheses. However, due to small size of the relationship in the stroke group, definitive conclusions regarding these relationships cannot be made. In other words, it is still possible that there is a stronger relationship between processing speed and loneliness in the stroke population than the healthy population, but we just did not have the right data to convincingly demonstrate the precise size of the effect.

The methodological approach employed in most prior work tends to be cross-sectional (Gilmour,2011; Gow et al. 2013; O’Luanaigh et al., 2012; Sin, Shoa, & Lee, 2021; Boss et al., 2015). It is worth noting that in cross-sectional research, the observed associations between variables may not necessarily imply causation as there is a possibility of confounding variables or shared underlying causes influencing the association, as demonstrated by Gow et al. (2013). Gow et al. (2013) initially identified a significant association between loneliness and processing speed. However, to clarify the nature of this relationship, they expanded their model to include depression symptom scores as a covariate. This adjustment revealed the previously observed association between loneliness and processing speed to become non-significant. This outcome underscores the importance of considering and controlling for potential confounding factors, or shared underlying causes, when interpreting findings from cross-sectional studies, as such factors can substantially impact the observed relationships between variables. It emphasises the need for caution in drawing causal conclusions solely based on cross-sectional data and highlights the value of employing covariates to enhance the precision and validity of study findings.

While the current study’s cross-sectional analysis revealed a significant association between loneliness and an increased incidence of stroke, the possibility of bidirectionality in this relationship should be acknowledged. Specifically, it is plausible that loneliness may contribute to a heightened risk of stroke, as suggested by the higher incidence observed among individuals with a single relationship status in the stroke group. However, it is equally important to consider the converse possibility – that individuals who have experienced a stroke may subsequently have faced increased challenges in maintaining social connections, leading to heightened feelings of loneliness. Likewise, individuals facing adversities such as heightened levels of socioeconomic deprivation may be more susceptible to both stroke and loneliness. To disentangle these complex dynamics, future longitudinal studies are warranted, allowing for a more comprehensive understanding of the temporal sequence and causal pathways underlying the observed associations.

Age is pivotal differentiating factor between prior research examining this area and is of relevance for the interpretation of our findings. Former studies primarily featured an older adult sample, surpassing the age of 70 years (O’Luanaigh et al. 2012; Gow et al., 2013; Oakely and Deary, 2019) whereas our current study encompassed participants within a comparatively younger age range, spanning from 46 to 48 years. The samples used in Sin et al. (2021) and Gilmour (2011) were marginally younger, with participants averaging 65 years of age. Nonetheless, the age of the participants remained older than that of the current study’s sample. It maybe that the relationship between cognition, in particular processing speed, and loneliness becomes more pronounced with increasing age. However, there was no consistent finding between processing speed and loneliness at the ages of 73 (r = -.071, p = .206), 76 (r = -.219, p=.001) 79 (r = -.114, p = .096) in Oakely & Deary’s (2019) longitudinal study. The only significant, but small, relationship was found at age 76 (Oakley and Dreary, 2019).

Apart from age, there are other design-related factors that could account for the disparities in findings between the present study and prior research in this field. Notably, all studies investigating the relationship between processing speed and loneliness have utilised varying measurement instruments to gauge processing speed. Primarily, two subtests (Symbol Search and Digit-Symbol) of the Wechsler Adult Intelligence Scale-III (WAIS-III) were utilised by Oakley & Dreary (2019), Gow et al. (2013), and O’Luanaigh et al. (2012) to measure psychomotor processing speed. Gow et al (2013) and Oakley & Dreary (2019) also utilised additional computerised reaction and inspection timed tests. Sin et al (2021) used the Symbol Digits Modality Test (SDMT). Overall, while all types of assessments aim to measure aspects of processing speed, they do so in slightly different ways. For example, the subtests from the WAIS-III, such as Symbol Search and Digit-Symbol, involve cognitive processes related to symbol recognition, matching, and coding. In contrast, the Letter Cancellation Task typically requires participants to scan and mark specific letters within a grid of random letters. The cognitive demands and processing requirements are somewhat distinct between these types of tasks.

Likewise, there is variability in the instruments employed to measure loneliness across the studies. Most previous studies (Oakley & Dreary, 2019; Gow et al., 2013; O’Luanaigh et al., 2012; Gilmour, 2011) relied on a single-question approach rated via a Likert Scale to gauge levels of loneliness. In contrast, Sin et al. (2021) adopted a more comprehensive approach by utilising the UCLA Loneliness Scale, a validated and reliable instrument specifically designed for measuring loneliness. In the present study, we opted for an alternative strategy by employing three items from the WEMWEBS (Warwick-Edinburgh Mental Well-being Scale) as a proxy measure for loneliness. The preliminary analysis revealed that the three WEMWEBS items exhibited a robust positive correlation with the GJGS (De Jong Gierveld Loneliness Scale), a well-validated measure of loneliness. This alignment enhances the validity of our approach and suggests convergent validity between our proxy measure and an established loneliness assessment tool, although it is acknowledged that this analysis is not a substitute for formal psychometric validation. The variations in the measurement of loneliness across studies hold the potential to explain the differences in the results and outcomes.

A consequence of secondary data analysis is that the research methodology relating to measuring variables of interest were beyond control of the current study. For example, inclusion of other assessments such as performance validity tests were not able to be implemented. This also extends to the number of participants in each group of interest. A key limitation of this study is the absence of data on the specific location and frequency of strokes experienced by participants in the BCS dataset. These stroke characteristics can significantly affect cognitive outcomes, yet their absence restricts our ability to explore variations in cognitive performance within our sample. Additionally, the reliance on self-reported stroke diagnosis introduces potential limitations in terms of accuracy compared to confirmed diagnoses from medical professionals. It’s also crucial to recognise that despite standardised administration procedures, minor variations in test administration or scoring across different studies may introduce some degree of heterogeneity in the results. This consideration remains pertinent in the current study, although the BCS acknowledges in their technical documentation that tests were conducted in standardized environments to ensure consistency (Centre for Longitudinal Studies, 2019). In addition, cognitive testing often measures a multitude of cognitive domains making it difficult to isolate specific cognitive processes. For example, although the LCT is a well-recognised test of processing speed, it is also dependent of visuomotor ability, in addition to sustained attention (McCrea & Robinson, 2011). Therefore, poor performance on the test could be attributed to an impairment in a different cognitive or motor domain, rather than processing speed ability. For example, those with visual neglect, typically associated with right hemisphere strokes, would score low on the LCT due to attentional impairment, not slower processing speed. Similarly, it may be that adopting a more comprehensive measure of loneliness may be more sensitive to detect the association between loneliness and processing speed. This, in turn, may result in a stronger relationship between loneliness and processing speed, and therefore measures such as the DJGLS or the UCLA Loneliness Scale (Russell, 1996) should be adopted.

Despite certain inherent limitations associated with this approach, it is noteworthy that this data derived from the British Cohort Study (BCS) offers several advantageous strengths in terms of cost-effectiveness, time efficiency, and the potential for replication. It also affords access to a substantial nationally representative sample, which enhancing the generalisability of the research findings. In addition, to ensure good research practice, the hypotheses, design and analysis were pre-registered prior to completion of the data analysis, which we consider a major strength in our approach. The analysis pipeline, and the dataset, is also accessible to aid replication of the analysis.

The clinical implications for the current study reinforce that loneliness should be routinely considered in stroke care. Healthcare professionals working with stroke survivors should be aware of the heightened risk of loneliness among this population. Assessing, and addressing, loneliness as part of stroke rehabilitation and follow-up care could improve overall well-being and potentially aid in recovery. As the study suggests that cognitive processing speed alone may not be a strong predictor of loneliness, future interventions to mitigate loneliness should consider a broader range of factors beyond cognitive processing speed, such as social engagement, emotional well-being, and personal relationships. This further emphasises the need for further comprehensive assessment to understand the multifaceted nature of loneliness and consider various contributing factors, which include cognitive and social dimensions, to inform more effective interventions. Furthermore, since the relationship between processing speed and loneliness appears to be both subtle and complex, clinicians should adopt a person-centred approach. Tailoring interventions and support based on the individual’s unique circumstances, including their cognitive abilities and health status, is crucial.

## Data Availability

All code and summary data produced are available here: https://osf.io/rxgdy/
Raw data can be accessed here: www.ukdataservice.ac.uk

## Acknowledgments

This research was performed as part of an all-Wales Economic and Social Research Council (ESRC) Doctoral Training Centre PhD studentship (awarded to RR and RC, PhD student: CB).

